# Clinical characteristics of COVID-19 and the model for predicting the occurrence of critically ill patients: a retrospective cohort study

**DOI:** 10.1101/2020.08.13.20173799

**Authors:** Jing Ouyang, Xuefeng Shan, Xin Wang, Xue Zhang, Yaling Chen, Miaomiao Qi, Chao Xia, Dongqing Gu, Yaokai Chen, Ben Zhang

## Abstract

**Background:** The present study aim to comprehensively report the epidemiological and clinical characteristics of the COVID-19 patients and to develop a multi-feature fusion model for predicting the critical ill probability.

**Methods:** It was a retrospective cohort study that incorporating the laboratory-confirmed COVID-19 patients in the Chongqing Public Health Medical Center. The prediction model was constructed with least absolute shrinkage and selection operator (LASSO) logistic regression method and the model was further tested in the validation cohort. The performance was evaluated by the receiver operating curve (ROC), calibration curve and decision curve analysis (DCA).

**Results:** A total of 217 patients were included in the study. During the treatment, 34 patients were admitted to intensive care unit (ICU) and no developed death. A model incorporating the demographic and clinical characteristics, imaging features and laboratory findings were constructed to predict the critical ill probability and it was proved to have good calibration, discrimination ability and clinic use.

**Conclusions:** The prevalence of critical ill was relatively high and the model may help the clinicians to identify the patients with high risk for developing the critical ill, thus to conduct timely and targeted treatment to reduce the mortality rate.

## 1. Introduction

In December 2019, the Health Commission of Hubei province firstly reported a group of unknown cause pneumonia patients relating to the South China seafood market in Wuhan city [1]. The novel coronavirus, named severe acute respiratory syndrome 2 (SARS-CoV-2) was quickly isolated and the virus-infected pneumonia was later designated as coronavirus disease 2019 (COVID-19) by The World Health Organization (WHO) [2]. Since then, the number of patients has increased rapidly, with confirmed cases successively appearing in many Chinese provinces (municipalities, autonomous regions and special administrative regions) and other 147 countries, areas or territories [3, 4]. As of August 1st, 2020, the globally reported laboratory-confirmed cases were nearly 1,900 thousands and WHO has declared COVID-19 as an international pandemic public health emergency[3, 5].

Up to now, a series of studies try to explain the clinical characteristics of COVID-19 and the clinical spectrum was reported to range from asymptomatic infection to severe pneumonia that should be admitted to the intensive care unit (ICU) and even death [6-9]. With the emergence of the second or third-generation infection of COVID-19, more clinical characteristics in other areas besides Wuhan need to be reported. Chongqing has 40 districts under its jurisdiction, being the largest municipality in China that adjacent to Wuhan. Until now, a total of 583 patients were diagnosed in Chongqing, but the clinical spectrums were not reported before[4].

Most of the COVID-19 patients were reported to have mild symptoms and a relatively better prognosis, while the critical ill patients hold poor prognosis and pose high risk for death [9, 10]. There is no specialized medicine to cure COVID-19 and the supportive care became the mainstay treatment regimen [11]. Therefore, to early identify the patients with high risk for developing critical ill and provide targeted care and treatment may prevent the patients from progression and reduce mortality risk, however, the valid tools are lacking.

Thus, in the present study, we aim to describe the epidemiological and clinical characteristics of COVID-19 patients who were diagnosed in the Chongqing city and then to construct and validate a nomogram for predicting the risk for developing critical ill patients, to help the clinicians have an early identification of the high risk patients and tailor targeted treatment regimens and reduce the mortality risk.

## 2. Materials and Methods

### 2.1. Study design and Participants

It was a retrospective cohort study. All of the COVID-19 patients who were diagnosed in the Chongqing Public Health Medical Center (Chongqing, China) between 24^th^ January and 16^th^ February 2020 were included and the data cutoff for the follow-up was 9^th^ March, 2020. The patients who were laboratory diagnosed according to WHO interim guidance in urban and surrounding areas of Chongqing were admitted to this hospital without selectivity[12].

For the development of the model for predicting the critical ill patients, all of the included patients were randomly splitting into two cohorts, namely the construction cohort (70%) and validation cohort (30%). The study was approved by the institutional review board of the Chongqing Public Health Medical Center (2020-017-01-KY).

### 2.2. Data sources

The demographic and epidemiological characteristics such as age, gender, past medical history, exposure history to Wuhan or the infected patients were collected by one professional personnel using a structured face-to-face questionnaire. The clinical characteristics such as the symptoms, imaging features, laboratory findings, treatment regimens, comorbidities and outcomes (under treatment, discharged or dead) for all of the patients were retrospectively reviewed and extracted from the electronic medical record using a unified data collection form.

### 2.3. Factors and Outcome definition

The primary end point of the present study was the critical ill pneumonia, which was defined as the COVID-19 patients who were admitted to the intensive care unit (ICU) required mechanical ventilation or have ≥ 60% inspired oxygen (FiO2) [11]. Comorbidities of acute kidney injury was defined based on the highest serum creatinine level and urine output[13]. The incubation period was defined as the interval between the earliest date of exposure to the Wuhan or the contact of the infected patients and the earliest date of symptom onset. For the patients who were usually worked in Wuhan, the latest date of exposure was considered as the time of infection[8]. Acute respiratory distress syndrome (ARDS) were defined according to the WHO interim guidance[12]. Secondary infection was made basing on the occurrence of symptoms or sign of hospital-acquired new pathogen infection after admission [9], and blood sepsis was defined as life-threatening organ dysfunction caused by infection according to the Third International Consensus Definition for Sepsis and Septic Shock (Sepsis-3)[14]. Recurrent COVID-19 was defined as the patients hospitalized again due to a positive test after discharge.

### 2.4. Statistical analysis

For the construction of the model for predicting the occurrence of the critical ill, the features with *p* <0.1 in the construction cohort were further analysed by using the least absolute shrinkage and selection operator (LASSO) method for dimensionality reduction and feature selection[15]. A nomogram was constructed basing on the logistic regression analysis to provide a much more understandable measure. The calibration ability of the nomogram was evaluated by the calibration curve. The discrimination ability of the nomogram was assessed by the area under the receiver operating curve (AUC). The difference in the AUC estimates was compared by Delong non-parametric test[16]. The Decision curve analysis (DCA) was additionally applied to calculate the net benefit by using the nomogram at different threshold probabilities[17].

Statistical analyses were conducted using Statistical Package for the Social Sciences (SPSS) version 23.0 software package for Windows (SPSS Inc), and R version 3.4.1 (R Foundation for Statistical Computing, Vienna, Austria; www.r-project.org). Statistically significant levels were two-tailed and set at *P*<0.05.

## 3. Results

### 3.1. Demographic and epidemiologic characteristics

A total of 217 patients were finally included in the current study, most of them were lived Chongqing city (N=208), seven from Hubei province and two from Sichuan province. (Figure 1) For these patients, 30.9% have exposure history to Wuhan, and 48.8% have contact history to infected patients (Table 1, Figure 2A). Median period for the incubation and time interval form illness onset to hospital were 7.00±7.3 days and 4.5±5.0 days, respectively. (Figure 2B) About half of the patients have infected family members (N=102, 47.0%), and the median infected family members was 1.0±2.0. (Figure 2C)

**Figure 1:**
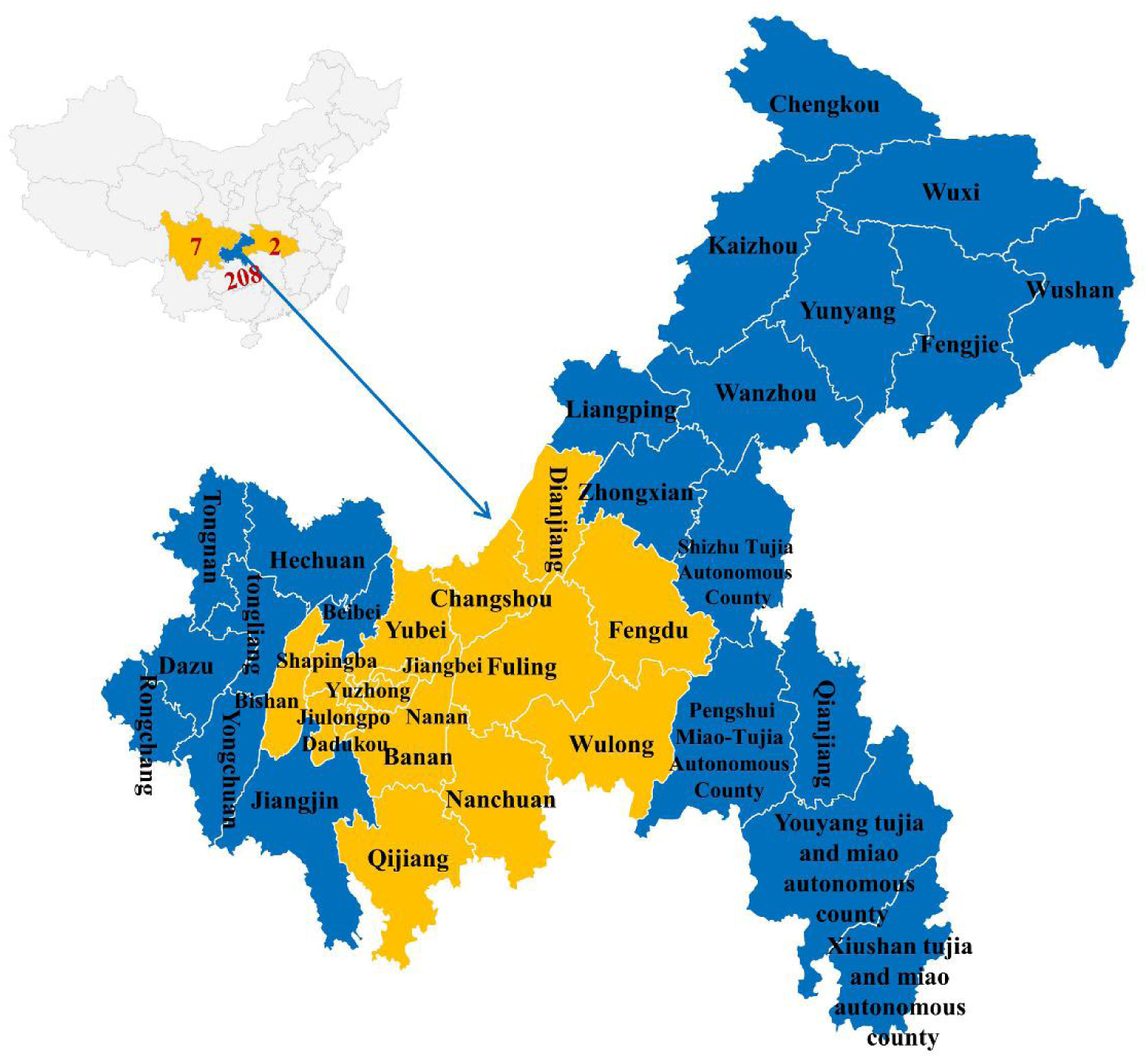
Geographical distribution of the included COVID-19 patients in the present study.

**Table 1.** Baseline demographic and clinical characteristics and outcomes in admitted to Chongqing Public Health Hospital.

| Character | Total cohort<br>(N=217, 100%) | Construction cohort<br>(N=159, 70%) | Validation cohort<br>(N=58, 30%) |
| --- | --- | --- | --- |
| **Demographic characteristics** |  |  |  |
| Age (mean ± SD, years) | 46.5±16.0 | 45.6±16.6 | 49.1±14.0 |
| Male | 101(46.5) | 73(45.9) | 28(48.3) |
| Married or divorce | 189(87.1) | 133(83.6) | 56(96.6) |
| Pregnant | 0(0.0) | 0(0.0) | 0(0.0) |
| Smoke | 36(16.6) | 28(17.6) | 8(13.8) |
| Drink | 18(8.3) | 12(7.5) | 6(10.3) |
| **Past history** |  |  |  |
| Cardiovascular disease | 68(31.3) | 49(30.8) | 19(32.8) |
| Digestive disease | 9(4.1) | 5(3.1) | 4(6.9) |
| Nervous system disease | 4(1.8) | 3(1.9) | 1(1.7) |
| Cancer | 1(0.5) | 1(0.6) | 0(0.0) |
| Diabetes disease | 23(10.6) | 18(11.3) | 5(8.6) |
| Asthma | 1(0.5) | 0(0.0) | 1(1.7) |
| COPD | 0(0.0) | 0(0.0) | 0(0.0) |
| **Clinical symptoms** |  |  |  |
| Fever ($\geq 37.3^{\circ}\text{C}$) | 107(49.3) | 75(47.2) | 32(55.2) |
| Cough | 140(64.5) | 107(67.3) | 33(56.9) |
| Expectoration | 75(34.6) | 56(35.2) | 19(32.8) |
| Shortness of breath | 29(13.4) | 23(14.5) | 6(10.3) |
| Muscular soreness | 26(12.0) | 12(7.5) | 14(24.1) |
| Unconsciousness | 0(0.0) | 0(0.0) | 0(0.0) |
| Headache | 23(10.6) | 15(9.4) | 8(13.8) |
| Dizziness | 23(10.6) | 15(9.4) | 8(13.8) |
| Poor appetite | 16(7.4) | 11(6.9) | 5(8.6) |
| Pharyngeal pain and discomfort | 29(13.4) | 24(15.1) | 5(8.6) |
| Chest pain | 3(1.4) | 3(1.9) | 0(0.0) |
| Nausea and vomiting | 7(3.2) | 5(3.1) | 2(3.4) |
| Running nose | 13(6.0) | 9(5.7) | 4(6.9) |
| Diarrhea | 19(8.8) | 13(8.2) | 6(10.3) |
| Fatigue | 48(22.1) | 30(18.9) | 18(31.0) |
| Chilly | 25(11.5) | 16(10.1) | 9(15.5) |
| Nasal obstruction | 8(3.7) | 5(3.1) | 3(5.2) |
| **Imaging feature** |  |  |  |
| Pulmonary infection |  |  |  |
| Unilateral infiltration | 8(3.7) | 8(5.0) | 0(0.0) |
| Bilateral infiltration | 207(95.4) | 149(93.7) | 58(100.0) |
| Ground-glass opacity | 180(82.9) | 128(80.5) | 52(89.7) |
| Pleural thickening | 111(51.2) | 78(49.1) | 33(56.9) |
| Pleural effusion | 6(2.8) | 4(2.5) | 2(3.4) |
| **Laboratory findings** |  |  |  |
| Blood glucose (mmol/L) |  |  |  |
| <6.1 | 117(53.9) | 85(53.5) | 32(55.2) |
| 6.1-7.0 | 65(30.0) | 47(29.6) | 18(31.0) |
| >7.0 | 35(16.1) | 27(17.0) | 8(13.8) |
| White blood cell (Per10<sup>9</sup>/L) |  |  |  |
| <4 | 62(28.6) | 41(25.8) | 21(36.2) |
| 4-19 | 148(68.2) | 114(71.7) | 34(58.6) |
| >10 | 7(3.2) | 4(2.5) | 3(5.2) |
| Lymphocyte count (Per10<sup>9</sup>/L) |  |  |  |
| <1.1 | 66(30.4) | 46(28.9) | 20(34.5) |
| 1.1-3.2 | 149(68.7) | 111(69.8) | 38(65.5) |
| >3.2 | 2(0.9) | 2(1.3) | 0(0.0) |
| Platelets (Per10<sup>9</sup>/L) |  |  |  |
| <125 | 34(15.7) | 24(15.1) | 10(17.2) |
| 125-350 | 173(79.7) | 127(79.9) | 46(79.3) |
| >350 | 10(4.6) | 8(5.0) | 2(3.4) |
| Hemoglobin (g/L) |  |  |  |
| <130 | 82(37.8) | 59(37.1) | 23(39.7) |
| 130-175 | 135(62.2) | 100(62.9) | 35(60.3) |
| D-dimer (ug/L) |  |  |  |
| ≤ 0.5 | 177(81.6) | 128(80.5) | 49(84.5) |
| 0.5-1.0 | 30(13.8) | 24(15.1) | 6(10.3) |
| >1.0 | 10(4.6) | 7(4.4) | 3(5.2) |
| C-reactive protein (mg/L) |  |  |  |
| 0.0-5.0 | 89(41.0) | 67(42.1) | 22(37.9) |
| >5.0 | 128(59.0) | 92(57.9) | 36(62.1) |
| **Treatment** |  |  |  |
| Oxygen therapy | 148(68.2) | 110(69.2) | 38(65.5) |
| Antiviral therapy | 214(98.6) | 156(98.1) | 58(100.0) |
| Interferon therapy | 216(99.5) | 159(100.0) | 57(98.3) |
| Antibiotic therapy | 50(23.0) | 36(22.6) | 14(24.1) |
| Antifungal therapy | 2(0.9) | 1(0.6) | 1(1.7) |
| Glucocorticoid therapy | 19(8.8) | 10(6.3) | 9(15.5) |
| Immunoglobulin therapy | 8(3.7) | 6(3.8) | 2(3.4) |
| Chinese herbs treatment | 114(52.5) | 84(52.8) | 30(51.7) |
| Protect liver treatment | 58(26.7) | 46(28.9) | 12(20.7) |
| Nutrition therapy | 15(6.9) | 11(6.9) | 4(6.9) |
| **Comorbidities and Outcomes** |  |  |  |
| Acute kidney injury | 2(0.9) | 0(0.0) | 29(3.4) |
| Liver injury | 71(32.7) | 52(32.7) | 19(32.8) |
| ARDS | 20(9.2) | 14(8.8) | 6(10.3) |
| Secondary infection | 0(0.0) | 0(0.0) | 0(0.0) |
| Blood sepsis | 5(2.3) | 2(1.3) | 3(5.2) |
| Admitted to ICU | 34(15.7) | 25(15.7) | 9(15.5) |
| Admission to discharge time<br>(median± quartiles, days) | 19.0±14.0 | 19.0±14.0 | 19.5±14.0 |
| Treatment outcome |  |  |  |
| Under treatment | 78(35.9) | 61(38.4) | 17(29.3) |
| Hospital discharge | 139(64.1) | 98(61.6) | 41(70.7) |
| Death | 0(0.0) | 0(0.0) | 0(0.0) |
| Recurrent SARS-Cov-2 | 5 (2.3) | 4(2.5) | 1(1.7) |

**Abbreviations:** COPD: chronic obstructive pulmonary disease; ARDS: Acute respiratory distress syndrome; ICU: Intensive Care Unit.

**Figure 2:**
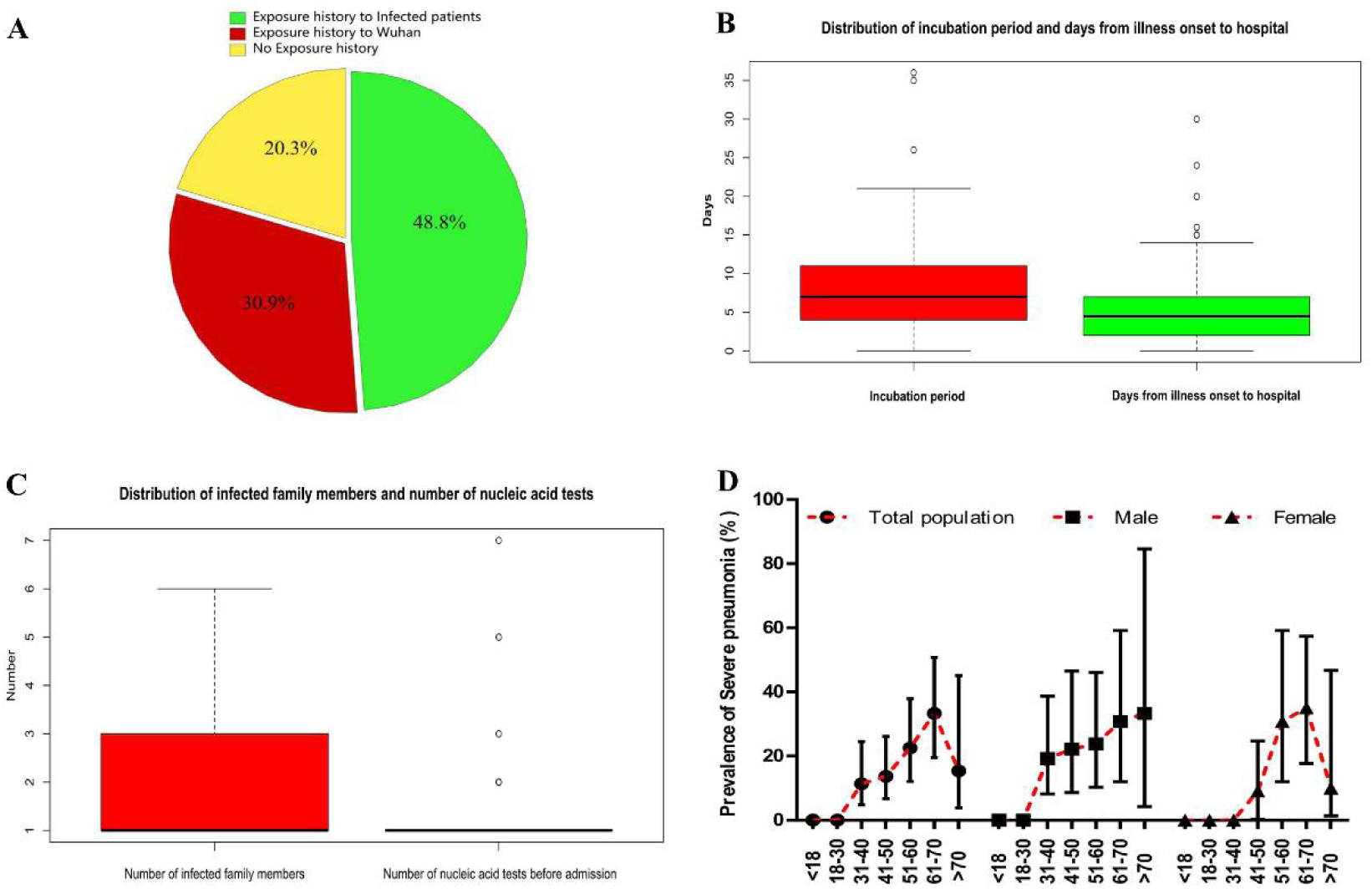
Distribution of the epidemiological characteristics of the included COVID-19 patients. A: Proportion of exposure history of included patients; B: Incubation and time interval between illness and hospital admission distribution; C: Number of infected family member and nucleic acid test before admission of included patients; D: The age and sex distribution of the critical ill patients.

### 3.2. Clinical characteristics

As shown in Table 1, the cardiovascular disease was the most common comorbidities for these patients (31.1%), followed by diabetes mellitus (10.6%), and digestive disease (4.1%). At admission, the most common symptoms were cough (64.5%) and fever (49.3%) Almost all of the patients have bilateral infiltration pulmonary infection (95.4%). For the laboratory findings, the leukopenia and lymphocytopenia occurred in 28.6% and 30.4% of the patients, respectively. Most of the patients received interferon therapy (99.5%) and antiviral therapy (98.6%). During the treatment, 32.7% presented liver injury, and 9.2% have acute respiratory distress syndrome (ARDS). The median time interval between admission and discharge time was 19.0±14.0 days. At the end of follow-up, six patients (4.3%) experienced recurrent COVID-19 after discharge.

### 3.3. Prevalence of critical ill

A total of 34 critical ill patients (15.7%) were admitted to ICU. The prevalence of critical ill was significantly increased with age and peaked at 61-70 years (33.3%; 95%CI: 19.5%-50.8%) then decreased. (Figure 2D) The prevalence in male and females presented different trends with age, however no difference was found in the total critical ill prevalence between them (18.8% for male, 12.9% for female, *X*^2^=1.41; *p*=0.24). (Figure 2D)

### 3.4. Differences between the critical ill and other patients in construction cohort

A total of 159 participants with 25 critical ill patients were incorporated into the construction cohort. Results suggested the critical ill patients presented high proportion of advanced age, married or divorced status, diabetes mellitus, more symptoms of fever, expectoration, short of breath and fatigue, imaging presentation of ground-glass opacity, pleural thickening, pleural effusion and more patients with high blood glucose, and lymphocytopenia. (Table 2)

**Table 2:** Difference in the demographic and clinical characteristics between severe and non-severe pneumonia patients in the construction cohort.

| Characteristics | Critical ill<br>(N, %) | Non-critical ill<br>(N, %) | $\chi^2/Z$ | *p*-value |
| --- | --- | --- | --- | --- |
| **Demographic characteristics** |  |  |  |  |
| Age (years) |  |  | 2.94 | <0.01 |
| ≤ 40 | 4(16.0) | 59(44.0) |  |  |
| 41-60 | 12(48.0) | 53(39.6) |  |  |
| >60 | 9(36.0) | 22(16.4) |  |  |
| Sex, (Male) | 13(52.0) | 60(44.8) | 0.44 | 0.51 |
| Marriage (Married or divorce) | 25(100.0) | 108(80.6) | 5.80 | 0.02 |
| Smoke | 4(16.0) | 24(17.9) | 0.05 | 0.82 |
| Drink | 3(12.0) | 9(6.7) | 0.84 | 0.36 |
| Exposure to Wuhan | 9(37.5) | 38(28.4) | 0.81 | 0.37 |
| **Past history** |  |  |  |  |
| Cardiovascular disease | 8(32.0) | 41(30.6) | 0.02 | 0.89 |
| Digestive disease | 1(4.0) | 4(3.0) | 0.07 | 0.79 |
| Nervous system disease | 0(0.0) | 3(2.2) | 0.57 | 0.45 |
| Diabetes disease | 8(32.0) | 10(7.5) | 12.64 | <0.01 |
| **Clinical symptoms** |  |  |  |  |
| Fever (≥ 37.3°C) | 18(72.0) | 57(42.5) | 7.34 | 0.01 |
| Cough | 21(84.0) | 86(64.2) | 3.76 | 0.05 |
| Expectoration | 18(72.0) | 38(28.4) | 17.59 | <0.01 |
| Shortness of breath | 9(36.0) | 14(10.4) | 11.12 | <0.01 |
| Muscular soreness | 0(0.0) | 12(9.0) | 2.42 | 0.12 |
| Headache | 3(12.0) | 12(9.0) | 0.23 | 0.63 |
| Dizziness | 2(8.0) | 13(9.7) | 0.07 | 0.79 |
| Poor appetite | 3(12.0) | 8(6.0) | 1.19 | 0.28 |
| Pharyngeal pain and discomfort | 2(8.0) | 22(16.4) | 1.17 | 0.28 |
| Chest pain | 0(0.0) | 3(2.2) | 0.57 | 0.45 |
| Nausea and vomiting | 1(4.0) | 4(3.0) | 0.07 | 0.79 |
| Running nose | 1(4.0) | 8(6.0) | 0.15 | 0.70 |
| Diarrhea | 0(0.0) | 13(9.7) | 2.64 | 0.10 |
| Fatigue | 9(36.0) | 21(15.7) | 5.69 | 0.02 |
| Chilly | 4(16.0) | 12(9.0) | 1.16 | 0.28 |
| Nasal obstruction | 0(0.0) | 5(3.7) | 0.96 | 0.33 |
| **Imaging features** |  |  |  |  |
| Pulmonary infection |  |  | 1.60 | 0.21 |
| Unilateral infiltration | 0(0.0) | 8(6.1) |  |  |
| Bilateral infiltration | 25(100.0) | 124(93.9) |  |  |
| Ground-glass opacity | 25(100.0) | 103(76.9) | 7.18 | 0.01 |
| Pleural thickening | 20(80.0) | 58(43.3) | 11.37 | <0.01 |
| Pleural effusion | 3(12.0) | 1(0.7) | 10.88 | <0.01 |

| **Laboratory findings** |  |  |  |  |
| --- | --- | --- | --- | --- |
| Blood glucose (mmol/L) |  |  | 2.78 | 0.01 |
| <6.1 | 8(32.0) | 77(57.5) |  |  |
| 6.1-7.0 | 8(32.0) | 39(29.1) |  |  |
| >7.0 | 9(36.0) | 18(13.4) |  |  |
| White blood cell (Per10<sup>9</sup>/L) |  |  | 0.59 | 0.56 |
| <4 | 7(28.0) | 34(25.4) |  |  |
| 4-19 | 15(60.0) | 99(73.9) |  |  |
| >10 | 3(12.0) | 1(0.7) |  |  |
| Lymphocyte count (Per10<sup>9</sup>/L) |  |  | 3.72 | <0.01 |
| <1.1 | 15(60.0) | 31(23.1) |  |  |
| 1.1-3.2 | 10(40.0) | 101(75.4) |  |  |
| >3.2 | 0(0.0) | 2(1.5) |  |  |
| Plates (Per10<sup>9</sup>/L) |  |  | 0.17 | 0.87 |
| <125 | 5(20.0) | 19(14.2) |  |  |
| 125-350 | 17(68.0) | 110(82.1) |  |  |
| >350 | 3(12.0) | 5(3.7) |  |  |
| Hemoglobin (g/L) |  |  | 2.82 | 0.09 |
| <130 | 13(52.0) | 46(34.3) |  |  |
| 130-175 | 12(48.0) | 88(65.7) |  |  |
| C-reactive protein (mg/L) |  |  | 14.18 | <0.01 |
| 0.0-5.0 | 2(8.0) | 65(48.5) |  |  |
| >5.0 | 23(92.0) | 69(51.5) |  |  |
| D-dimer (ug/L) |  |  | 1.94 | 0.05 |
| ≤ 0.5 | 17(68.0) | 111(82.8) |  |  |
| 0.5-1.0 | 4(16.0) | 20(14.9) |  |  |
| >1.0 | 4(16.0) | 3(2.2) |  |  |

### 3.5. The model for predicting critical ill probability

A total of 13 factors (two for demographic factors, six for clinical factors, three for imaging features and two for laboratory findings) were finally selected. (Figure 3A-B) And then a nomogram was constructed for predicting the critical ill occurrence probability. (Figure 3C)

**Figure 3:**
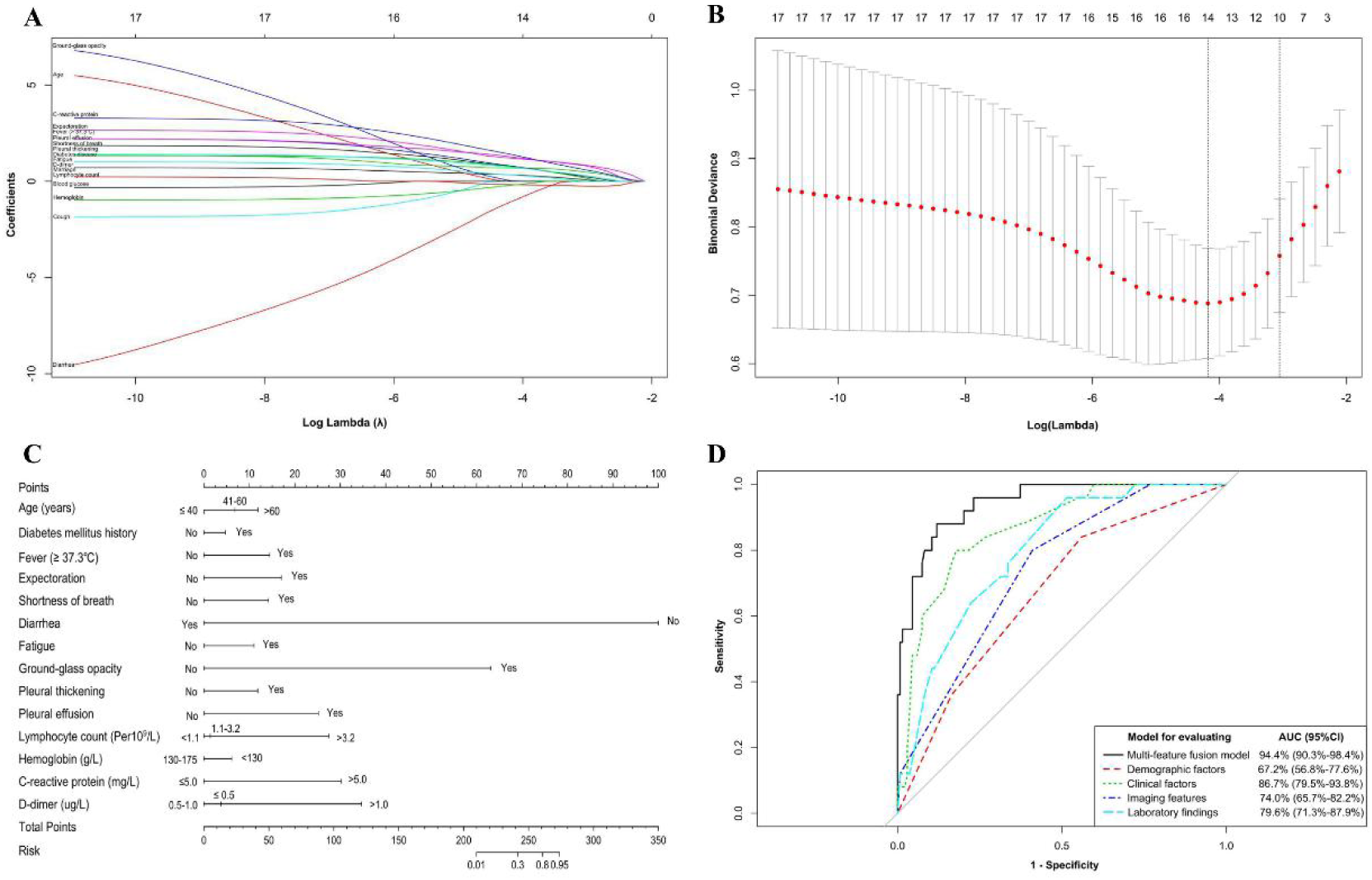
Factors selection by the least absolute shrinkage and selection operator (LASSO), the model construction and the comparison of the discrimination among different models. A: The LASSO coefficient profile of the thirteen selected factors; B: Selection of the tuning parameter (λ) in the LASSO model using 10-fold cross-validation via binomial deviance minimization criteria. C: The nomogram for predicting the probabilities of critical ill occurrence; D: The comparison of the AUC among different models for predicting critical ill occurrence.

The AUC for the predicting nomogram was 92.2% (95%CI: 87.8%-96.6%). (Figure 3D) When comparing the performance among different prediction models, the clinical factors model presented comparable performance with the full model (*p*=0.06) and have significantly higher performance than demographic (*p*=0.01), imaging features (*p*=0.03) and laboratory findings model (*p*=0.02). The calibration curve suggested good performance of the nomogram. (Figure 4A) Decision curve analysis also suggested with the threshold probability is over 20%, the application of the nomogram for predict critical ill and conducted targeted treatment adds more benefit than treating all or none of the patients. (Figure 4B)

**Figure 4:**
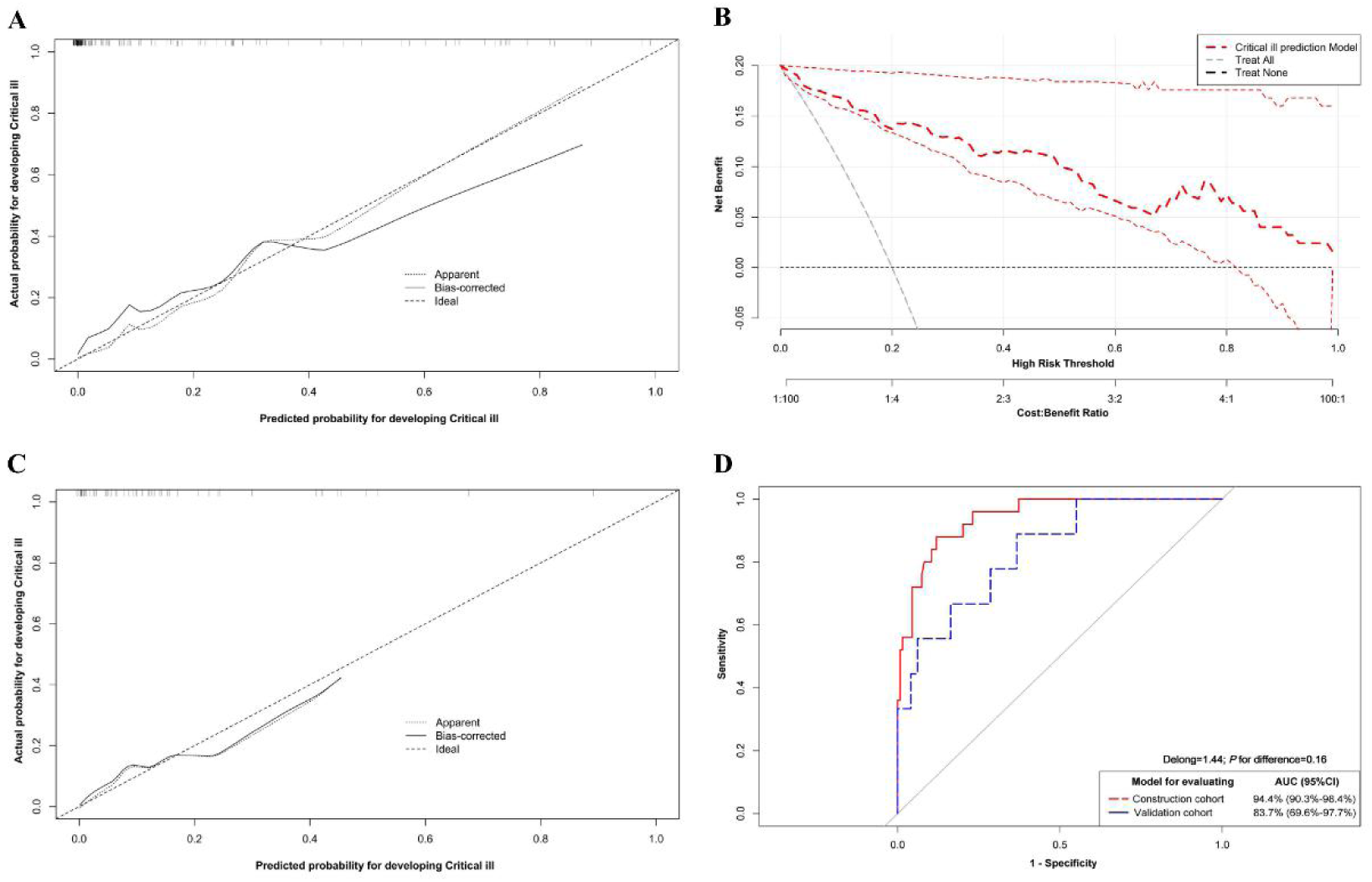
Validation of the performance, clinical use and transportability of the nomogram. A: The calibration curve for the nomogram in the construction cohort; B: Decision curve analysis of the net benefit by using the prediction nomogram in the construction cohort; C: The calibration curve of the nomogram in the validation cohort; D: The difference in the discrimination ability for nomogram between the construction and validation cohort.

When applying the nomogram in the validation cohort, the results showed relatively good agreement between predicted and observed critical ill probability with AUC of 87.3% (95%CI: 77.1%-97.5%), and no difference was found between the construction and validation cohort (92.2% versus 87.3%, D=0.87; *p*=0.39). (Figure 4C-D)

## 4. Discussion

The present study comprehensively described the epidemiological and clinical characteristics of COVID-19 in Chongqing. One third of the 217 patients have exposure history to Wuhan and another half reported to have contact with infected patients, which further confirmed the human to human transmission of the COVID-19 and indicated most of the included patient maybe the second or more generations of infection. Moreover, the family cluster outbreak make up the main source of patients in this study. About 47% of the patients were reported to have infected family members and the median number of infected family member was 1 (maximum 7 members), which confirm the previous family cluster reports [18] and further indicated the importance of family protection. Mean age of them were 46.5 years, nearly half of them were male and teenagers were seldom infected, which were similar to the findings from a national multicenter investigation[8]. However, the incubation period of the present study was relatively longer than before [8, 19, 20].

In line with the previous reports, the present study also suggested fever and cough were the most common symptoms at admission [6, 9, 10, 20]. Additionally we found COVID-19 may present chilly and poor appetite, which was seldom reported before. CT imaging feature also plays important roles on the identification of the COVID-19 patients as it is reported that the CT imaging findings predate the RT-PCR positive results [21, 22]. In this study, most of the patients present bilateral infiltration with ground-glass opacity on the CT scan, which was in line with the previous studies [6, 9, 10, 20]. Moreover, we also found pleural thickening in half of the patients and a small fraction of them presented pleural effusion, which was seldom reported at this time. In keeping with the finding in other studies, we also found part of the patients presented with leukopenia, lymphocytopenia, anemia and thrombocytopenia; however, due to the sample size and distribution of the clinical spectrum of the patients, the proportions were not consistent [7-9, 10, 20]. For the lack of the specific treatment, all of the patients underwent supportive care. A lot of marketed and unmarketed drugs and Chinese herbs such as Remdesivir and Lianhua qingwen capsule are in clinical trials; the curative effect will be revealed later and applied to the clinic as soon as possible.

The results also showed the prevalence of critical ill was relatively smaller than the previous studies reported by Huang (31.7%) [9], Zhou (26%) [10] and Wang colleagues (26.1%)[6]. Moreover, all of the included patients in this study presented relatively smaller proportion of comorbidities and death than them. The phenomenon may be partly explained by the relatively smaller time interval from illness to admission of the present study [6, 9, 10]. Thus early diagnosis and have timely treatment of the infected patients might may benefit of the patients from high risk of critical ill progress, comorbidities occurrence and even death. In addition, although with low frequency, we also found 2.8% of the patients have recurrent COVID-19 after hospital discharge. It should be served as a reminder that patients who have been discharged from the hospital need to continue to be isolated and protected to some extent avoiding the recurrent and lead to transmission again.

Critical ill predispose patients high risk to death [10, 11]. Comparing with the controls, the present study showed the critical ill patient presented advanced age, high proportion of diabetes mellitus, short of breath, and lymphopenia, which was consistent with Wang et al[6]. However, Huang and colleagues did not suggest this difference, which may due to the relatively smaller sample size [9]. Additionally, we also found the critical ill patients present higher proportion of imaging findings of ground-glass opacity, pleural thickening and pleural effusion, which were seldom reported. It could be partly explained by the report that the changes in the chest CT imaging features were associated with the clinical manifestation from diagnosis to recovery and may predict the disease progression[21].

Basing on the aforementioned factors, the present study comprehensively developed a multi-feature fusion model to help the clinicians have early identification of the critical ill patients. After evaluation, the model was proved to have good internal and external performance and may provide important value for the medical decision making. Among these included factors, the clinical features showed a relatively better performance than the others, thus the symptoms and past disease history were the primary information for evaluating the severity of the patients. The demographic factors, imaging features and laboratory findings with comparable predicting value, were good supplement for the clinical characteristics. However, due to the limited sample size of patients, we cannot carry out clinical trials for validating its clinical application in this study.

Our study has some limitations. Firstly, the sample size incorporated into the research was relatively small, which may partly affect the statistic power of the results. Secondly, not all of the laboratory tests were done in all of the included patients such as D-dimer and proinflammatory cytokines, which were proved to play important roles in the critical ill occurrence [9]. But with all of the included features, the prediction model was proved to be have good performance and clinical use, thus they may not significantly affect the results.

To sum up, this study comprehensively describes the clinical characteristics of the COVID-19 patients and then to construct and validate a model for predicting the occurrence of critical ill probability. The results may help the clinicians to have an unrivalled understanding of the characteristics of COVID-19 patients and then to stratified the patients into different risk subgroups for developing critical ill and tailor targeted treatment regimens for the high risk patients and reduce the mortality rate.

## Data Availability

The availability of all data referred to in the manuscript.

## Acknowledgments

This work was supported by National Natural Science Foundation of China (81673255, 81874283, 81903398); Chongqing Special Research Project for Prevention and Control of Novel Coronavirus Pneumonia (No.cstc2020jscx-fyzx0074); The Recruitment Program for Young Professionals of China (No number); Funds from Army Medical University and First Affiliated Hospital of Army Medical University (2018XLC1004, SWH2018BJKJ-12). Chongqing Natural Science Foundation Program (cstc2019jcyj-msxmX0466).

## Author Contributions

Drs Jing Ouyang, Xuefeng Shan, and Xin Wang had full access to all of the data in the study and take responsibility for the integrity of the data and the accuracy of the data analysis. Drs Ben Zhang, Yaokai Chen contributed equally as senior authors. Concept and design: Ben Zhang, Yaokai Chen; Acquisition, analysis, or interpretation of data: Xuefeng Shan, Xin Wang, Jing Ouyang, Xue Zhang, Miaomiao Qi, Chao Xia, Yaling Chen; Drafting of the manuscript: Xuefeng Shan, Xin Wang, Jing Ouyang; Critical revision of the manuscript for important intellectual content: Ben Zhang, Yaokai Chen; Statistical analysis: Xin Wang, Xue Zhang, Yaling Chen; Supervision: Ben Zhang, Yaokai Chen.

